# Proteome sampling with e-biopsy enables differentiation between cutaneous squamous cell carcinoma and basal cell carcinoma

**DOI:** 10.1101/2022.12.22.22283845

**Authors:** Edward Vitkin, Julia Wise, Ariel Berl, Ofir Shir-az, Batel Gabay, Amrita Singh, Vladimir Kravtsov, Zohar Yakhini, Avshalom Shalom, Alexander Golberg

## Abstract

Clinical misclassification between cutaneous squamous cell carcinoma (cSCC) and basal cell carcinoma (BCC) affects treatment plans and carries risks of potential for recurrence, metastases morbidity and mortality. We report the development of a novel tissue sampling approach with molecular biopsy using electroporation. The methods, coined e-biopsy, enables non-thermal permeabilization of cells in the skin for efficient vacuum-assistant extraction of informative biomolecules for rapid diagnosis. We used e-biopsy for *ex vivo* proteome extraction from 3 locations per patient in 21 cSCC and 21 BCC pathologically validated human tissue samples. The total 126 extracted proteomes were profiled using LC/MS/MS. The obtained mass spectra presented significantly different proteome profiles for cSCC and BCC with several hundreds of proteins significantly differentially expressed in each tumor in comparison to the other. Notably, 17 proteins were uniquely expressed in BCC and 7 were uniquely expressed in cSCC patients. Statistical analysis of differentially expressed proteins found 31 cellular processes, 23 cellular functions and 10 cellular components significantly different between cSCC and BCC. Machine Learning classification models constructed on the sampled proteomes enabled the separation of cSCC patients from BCC with average cross-validation accuracy of 81%, cSCC prediction positive predictive value (PPV) of 78.7% and sensitivity of 92.3%, which is comparable to initial diagnostics in a clinical setup. Finally, the protein-protein interaction analysis of the 11 most informative proteins, derived from Machine Learning framework, enabled detection of a novel protein-protein interaction network valuable for further understanding of skin tumors. Our results provide evidence that the e-biopsy approach could potentially be used as a tool to support cutaneous tumors classification with rapid molecular profiling.

## Introduction

Cutaneous squamous cell carcinoma (cSCC) and basal cell carcinoma (BCC) are heterogeneous skin lesions, which belong to a broad group of nonmelanoma skin cancer (NMSC)^1^. Both are increasing in incidence worldwide from 3 to 8% per year since the 1960’s^1^. cSCC, representing 20% of all skin cancers, accounts for approximately 75% of all skin cancer deaths, excluding melanoma^2^. BCC, which is the most common malignant neoplasm of humans, is usually curable when the lesion is treated in the early phase. It is estimated that 3.6 million cases of NMSC are diagnosed in the USA alone every year^3^, constituting an enormous financial burden on the healthcare systems^4,5^. Sun exposure is the most important risk factor for both cSCC and BCC. In addition, exposure to carcinogenetic agents, such as arsenic and aromatic hydrocarbons, and viral infections play an important causative association ^1^. Also, cSCC may arise in the setting of scars, draining sinuses, ulcers, burn sites, and areas of chronic inflammation^1^. The clinical diagnoses of cSCC and BCC are performed by a direct skin inspection^6^, often assisted by dermoscopy^7^. In addition, multiple emerging methods such as optical coherence tomography (OCT)^8^, reflectance confocal microscopy (RCM)^9^, elastic scattering spectroscopy (ESS)^10^ and high-frequency ultrasound (HFUS)^11^ aim to assist the clinician in diagnosis. Nevertheless, tissue biopsy for histopathological analysis is still essential to confirm the diagnosis, to estimate the risk of recurrence and to further dictate the treatment pathway^12^. Skin structural and functional complexity lead to existence of multiple types of biopsies, with major methods including punch biopsy, shave biopsy, excisional biopsy and curettage biopsies^1^.

The clinical features of cSCC and BCC are well described and, in most cases, the clinical diagnosis is done according to the subsequent histological verification. Sometimes, however, the clinical phenotypes of these cancers are ambiguous and discrepancies between the clinical presentations and histologic analyses occur ^13,14^. Such misclassification of cSCC as BCC (or vice versa) affects the treatment plan of the lesion^6^. Misclassification of cSCC as BCC carries the highest risk to patients, due to the inherent potential for recurrence, metastases and mortality^15^. Recent studies suggested that the introduction of molecular biomarkers, that would accurately stratify non-melanoma skin lesions, would provide a new frontier in personalization of skin cancer care, and hold a potential of greatly improving patient diagnosis in many cases ^16^.

Besides algorithms for discovery of the specific molecular signatures for each of the diseases, the yet open challenge is biomarkers sampling. The current strategy involves molecules extraction using lyses buffers from tissue sample with tissue biopsies. Tissue biopsy procedures carry risks involved with surgical procedures and may lead to localized tissue injury, bleeding, inflammation, infection and scarring^17^. In addition, because of these risks, only a few biopsies can be performed in a single procedure, limiting the scope of the spatial mapping of the entire lesion, potentially leading to misdiagnosis if the tumor is only partially sampled or completely missed^18^. To address these problems and to allow direct molecular sampling from tissue without resection, a series of procedures and associated mass spectrometric tools are currently under development^19^, yet none of these to date have received a broad community recognition. These include the intelligent knife (iKnife), desorption electrospray ionization (DESI)^20^, picosecond infrared laser (PIRL) and the MasSpec pen^19,21^. All these methods require high energy for sample evaporation, potentially damaging the tissue^19^. In addition, the ionization process is competitive, thus, not all informative molecules will be sampled^19^.

To address the need in improved tissue-spatial sampling of biomarkers, and to extend the technology state of the art in enabling precise personal medicine, we developed a novel tissue sampling approach with molecular biopsy using electroporation. Electroporation-based technologies have been successfully used to permeabilize the cell membrane *in vivo*, enabling a wide set of applications ranging from tumor ablation to targeted delivery of molecules to cell populations and tissues^22^. We have previously developed protocols for targeted delivery of electric fields to tissues to induce focused electroporation at predetermined regions in organs^23^. More recently, we have shown that electroporation-based molecular sampling, termed “e-biopsy”, selectively extracts liquids from solid tissues with informative proteomes in animal models in liver cancer^24^ and brain melanoma^25^ *in vitro* and from breast cancer *in vivo*, enabling *in vivo* spatial mapping of differential protein expression^26^. To the best of our knowledge, such a technology is not yet available and has not been reported in the literature for minimally invasive human tissue molecular sampling. Here we demonstrate the ability to sample and analyze the proteome extracted *ex vivo* by e-biopsy from excised human skin. Moreover, we show numerical models demonstrating a non-thermal nature of this technology together with the analysis of the sampling reproducibility.

This work tests the hypothesis that tissue liquids, sampled by e-biopsy, contain proteomic signatures relevant to skin tumors. Specifically, we show that proteomic profiles obtained by e-biopsy from cSCC are different from those obtained from BCC. We found proteins uniquely observed either in cSCC or in BCC and proteins significantly differentially expressed. Application of standard machine learning techniques allowed to identify a subset of proteins distinguishing between the two cancers with the classification performance comparable to current diagnostic methods that involve human professional inspection. Moreover, proteomic data produced by e-biopsy lead to a new protein-protein interaction network jointly covering many of these highly informative proteins – a finding that may potentially improve our understanding of systemic molecular differences between the two carcinomas and potentially create new target therapies.

Our novel e-biopsy approach to the characterization of skin tumors differs substantially from excision biopsy approaches, that require tissue resection and that provide information limited to the size and the region of resected tissue. E-biopsy is also different from direct mass spectrometric tools as it does not require tissue disrupting direct ionization but rather the use of non-thermal pulsed electric fields, which do not lead to skin damage. scarring or to other potential medical risks ^27–30^.

## Results

### Proteomics harvesting and analysis from excised human skin with e-biopsy

Electroporation-based biopsy (e-biopsy) for proteome sampling from freshly (10 to 20 minutes since the time of excision) excised human skin is shown in **Figure 1**. First, the sampling needle is inserted in the sampling location and the ground needle is positioned on the skin surface (without penetration) and the pulsed electric fields (PEF) are applied (**Figure 1a**). Second, a vacuum is applied on the same needle through which the PEF pulses are delivered, to pump the released cellular content into the needle and the syringe (**Figure 1b**). Next, the tissue extract (∼1-3µL) is discharged from the syringe to the external buffer (biology grade water), and subjected to standard protocols for molecular analysis, including purification, separation, identification (LC/MS/MS in this case), and quantification (**Figure 1c**). Next, statistical analysis was performed, and a machine learning classifier was constructed to determine a molecular signature that differentiates BCC and SCC tumors (**Figure 1d**). E-biopsy was repeated in three positions in the same area of the excised tissue sample.

**Figure 1.**
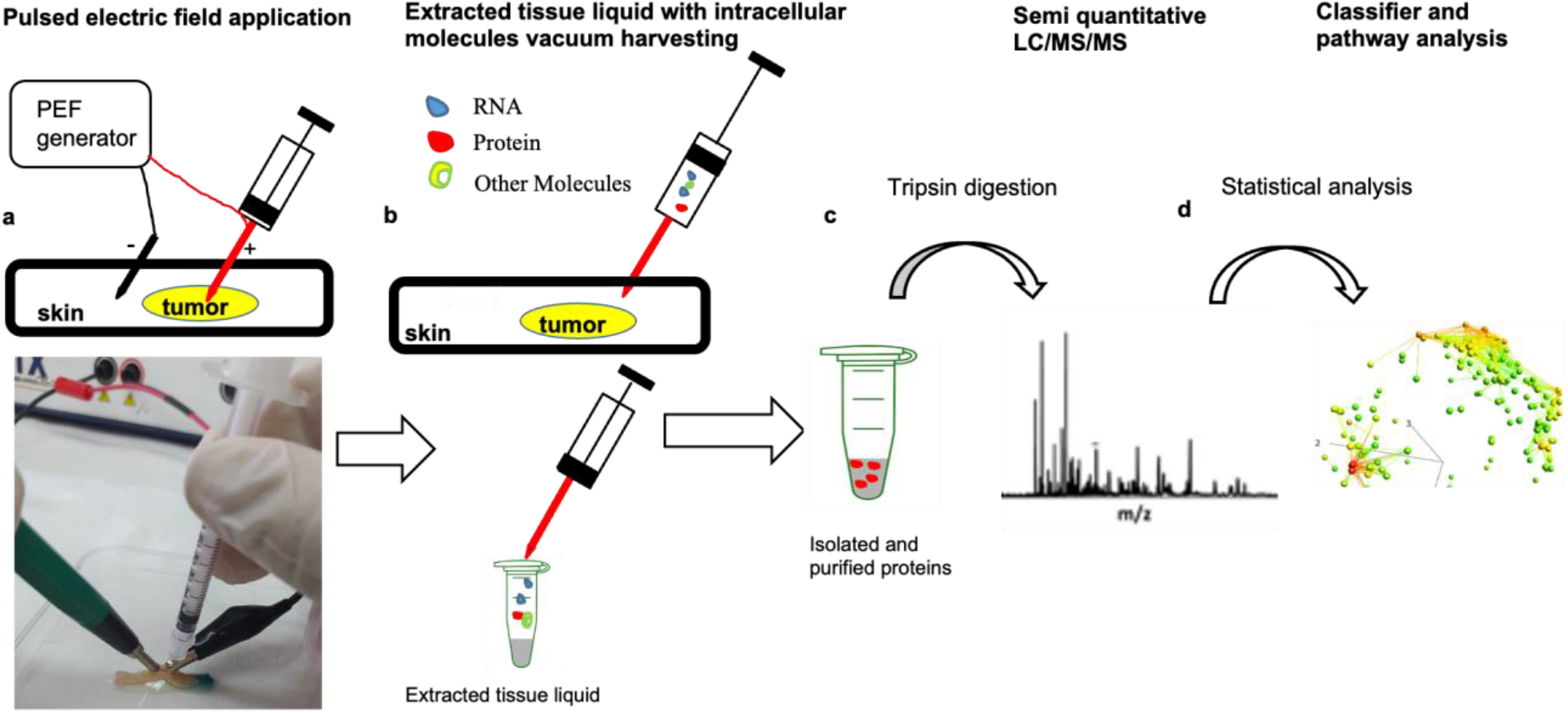
Skin e-biopsy procedure *ex vivo*. Schematics of molecular harvesting with e-biopsy (**a-d**). **a**. Application of high voltage pulsed electric fields on the excised skin. A sampling electrode (insulin syringe needle, connected to the voltage generator) is inserted inside the skin sample. The ground electrode is located on the skin surface. **b**. Sampling of the released and intercellular molecules applying vacuum to the needle. **c**. Isolation, purification and then identification and quantification of proteins, using shotgun proteomics. **d**. Bioinformatics analyses for differential expression and classifier construction.

### Modeling electric field in the human skin with tumors during e-biopsy

Numerical modeling is a computational tool that allows prediction of electric field and thermal distributions in physiologically complex structures. Usage of numerical models allows e-biopsy device designers to focus the electric fields in the specific sampling areas, limiting the potential damage to other tissues. We created a finite element model using a commercial Quick Field software (Tera Analysis Ltd, Denmark) to estimate the electric field and thermal distributions in the excised tissues, during e-biopsy. The model constitutes of a 2D mesh, in a plane-parallel mode and the problem type is direct current (DC) conduction. A general carcinoma lesion (the electrical^31,32^ and thermal^33–35^ properties of BCC and cSCC are similar) was modelled as a 2D rectangle with 20mm length, 2mm infiltration depth, and surrounded with a healthy skin strap of 5mm length from both sides of the lesion. Needle electrode with radius of 0.3mm is positioned 10mm from the ground electrode and it penetrates the skin 1mm into the lesion. Ground electrode is placed without skin penetration on the lesion surface area, with 3mm radius. The model mesh is presented in **Figure 2**:

**Figure 2.**
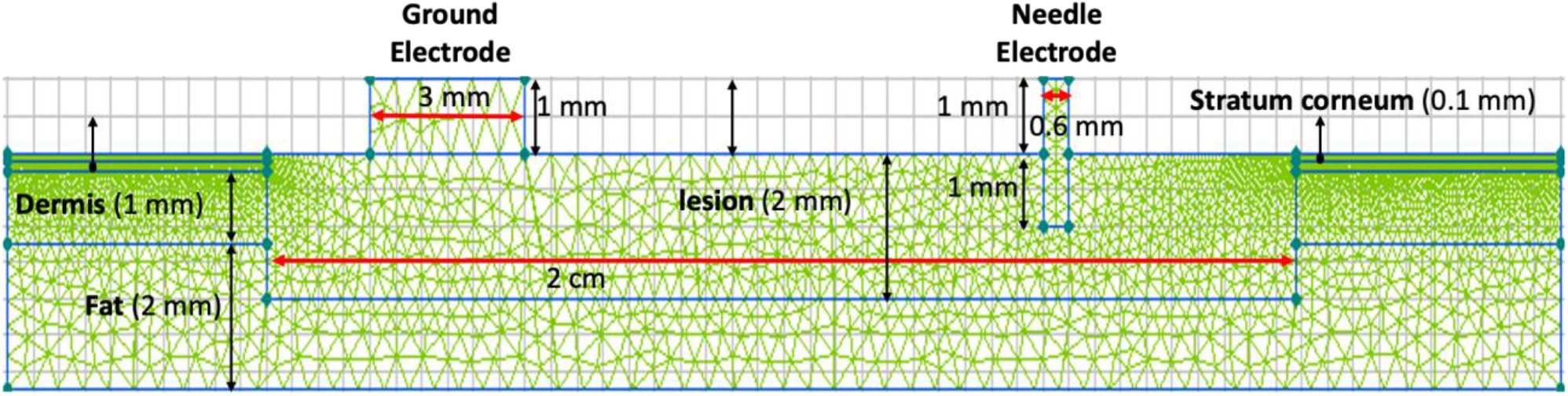
Model geometry. Skin layers are comprised of stratum corneum, epidermis and dermis, followed by fat layer, each layer has its own electrical properties, presented in **Table S1**. In this model a tumor penetrates 2mm, clear skin layers route into fat layer, and alongside lesion left and right boundaries, 5mm healthy skin tissue. Actual geometric proportions are not preserved here for visualization purposes.

It is crucial to know tissue properties for correct prediction of electroporation outcomes and of thermal effects. Electric field distribution is mainly determined by the electroporation protocol, such as pulse parameters, electrode configuration and physical features of the tissue. For simplicity, each layer in this model is assumed to have homogeneous and isotropic properties, taking into consideration the complexity of the skin layers as the stratum corneum, epidermis and dermis, all with anisotropic dielectric properties^36^, including lesion, that paved its way through skin layers, infiltrating fat layer. In this model (**Figure 3**), we selected the electric field strength of 480V cm^-1^ as a threshold for skin reversible electroporation^37^. The threshold selected for the irreversible skin electroporation was 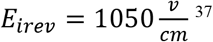. In addition, to estimate the potential thermal effects of the application of high-voltage pulsed electric fields on a skin tissue, we coupled the DC model with the transient heat field problem (**Figure 3**). The maximum predicted temperature increase was 2.2°C, reaching 27.2°C relative to the initial temperature of 25. 0°C (black dot on **Figure 3c**).

**Figure 3.**
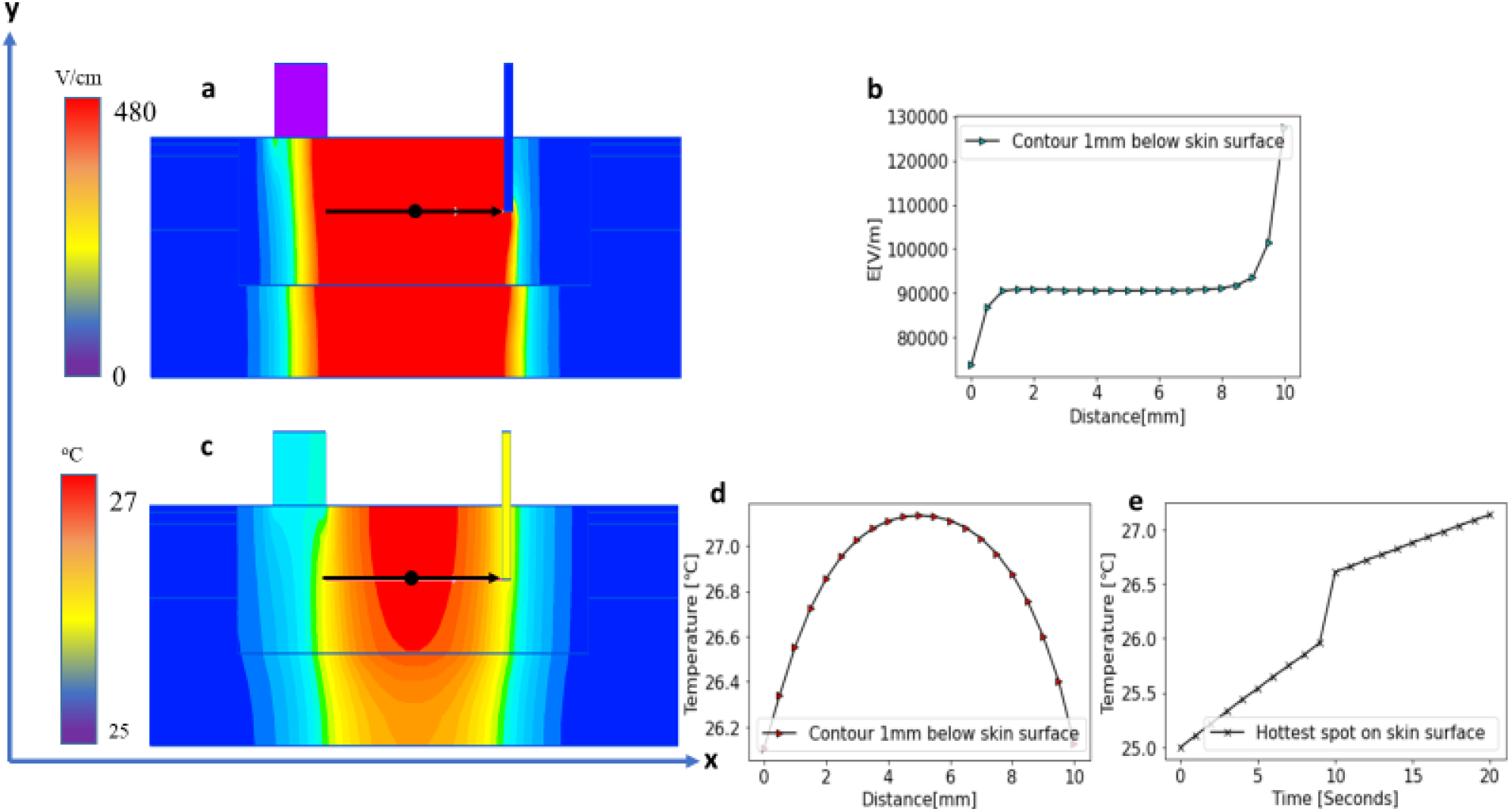
**(a)** The electric field distribution in a lesion while applying 1000V, with a color map of the electric field strength. Red color represents the electroporated tissue area (E>480V cm^-1^), which is mainly located between ground and needle electrodes. **(b**) An electric field strength profile between ground electrode to needle electrode, 1mm below lesion surface **(c**) Analysis of temperature distribution after two sequential pulses, high and rapid followed by slow and longer pulses. Transient heat transfer problem created through coupling of DC conduction problem when V=1000SV were applied, V_rms_ = 12.65V, pulsing for 10 seconds, with steady state heat transfer problem with confined Dirichlet boundary conditions. The described transient heat problem was coupled after 10 seconds pulsing, with another DC conduction problem when V=50V were applied, V_rms_ = 10V, pulsing for 10 seconds to create a transient heat transfer problem performed in a and d figures. **(d)** The temperature profile from the ground electrode until the needle electrode. **e** Temperature increase in time at the point located 1mm beneath the electrode.

### Reproducibility of molecular harvesting with e-biopsy in cutaneous SCC and in BCC

To assess the reproducibility of the e-biopsy methodology, the similarity between the measurements gathered from 3 measured for each patient was estimated. Our assumption was that the actual proteomes in the sampled locations should be very similar, given these locations are spatially and phenotypically close. Therefore, we expected that protein profiles sampled by a reliable technology to be in a high agreement with each other. Together with this, inherent tissue spatial heterogeneity would prevent even the ideal sampling method from receiving the exact measurement replicate.

Specifically, we calculated maximal intra-patient Pearson correlation between proteomes measured in each sampled patient’s location. We observed Pearson correlation of 0.914±0.058 (0.925 ± 0.052 for BCC patients; 0.903 ± 0.062 for SCC patients; **Table S2**), which indicates high consistency of the e-biopsy sampling technique.

### E-biopsy sampled proteome differentiates cutaneous SCC from BCC in human skin

To assess the data potential in distinguishing between the cSCC and the BCC proteomic profiles, we used a balanced dataset of 42 patients, specifically 21 cSCC and 21 BCC patients with similar gender, age, and birth origin distributions (**Figure S1**). Each patient’s lesion was sampled in 3 locations, resulting in 63 samples for each carcinoma type. In total, we identified 7087 proteins observed with non-zero intensity in at least one of 126 dataset samples.

The e-biopsy extracted proteomics profiles obtained from cSCC and BCC appeared notably different. Among all 7087 observed proteins we identified 118 proteins (FDR=1.9e-01, **Methods**), that completely do not appear (zero intensity) in cSCC, while appearing in at least 5% (at least 4) of BCC samples. Moreover, 17 of these proteins (FDR=9.8e-02) appear in at least 33% (at least 7 out of 21) of BCC patients (**Table 1, Table S3**). In addition, we identified 100 proteins (FDR=2.3e-01), that do not appear at all in BCC, while appearing in at least 5% (at least 4) of cSCC samples; 7 of them (FDR=2.4e-01) were identified in more than 33% (at least 7 out of 21) of cSCC patients (**Table 2, Table S4**).

**Table 1.** 17 most frequent genes unique to BCC samples compared to cSCC. Full table see **Table S3**

| GeneID | Frequency in samples # (%) | Frequency in patients # (%) | Gene Name |
| --- | --- | --- | --- |
| Q96I15 | 15(23.8%) | 9(42.9%) | Selenocysteine lyase;SCLY |
| Q6UXV4 | 13(20.6%) | 8(38.1%) | MICOS complex subunit MIC27;APOOL |
| P09132 | 11(17.5%) | 8(38.1%) | Signal recognition particle 19 kDa protein;SRP19 |
| Q16773 | 11(17.5%) | 8(38.1%) | Kynurenine--oxoglutarate transaminase 1;KYAT1 |
| P29972 | 10(15.9%) | 8(38.1%) | Aquaporin-1;AQP1 |
| Q13144 | 9(14.3%) | 8(38.1%) | Translation initiation factor eIF-2B subunit epsilon;EIF2B5 |
| P36954 | 9(14.3%) | 8(38.1%) | DNA-directed RNA polymerase II subunit RPB9;POLR2I |
| P23443 | 8(12.7%) | 8(38.1%) | Ribosomal protein S6 kinase beta-1;RPS6KB1 |
| P40967 | 11(17.5%) | 7(33.3%) | Melanocyte protein PMEL;PMEL |
| P12694 | 11(17.5%) | 7(33.3%) | 2-oxoisovalerate dehydrogenase subunit alpha, mitochondrial;BCKDHA |
| Q6PD62 | 11(17.5%) | 7(33.3%) | RNA polymerase-associated protein CTR9 homolog;CTR9 |
| A4D1P6 | 11(17.5%) | 7(33.3%) | WD repeat-containing protein 91;WDR91 |
| Q9Y3R5 | 10(15.9%) | 7(33.3%) | Protein dopey-2;DOP1B |
| Q9NP81 | 9(14.3%) | 7(33.3%) | Serine--tRNA ligase, mitochondrial;SARS2 |
| Q92990 | 9(14.3%) | 7(33.3%) | Glomulin;GLMN |
| Q8N7H5 | 9(14.3%) | 7(33.3%) | RNA polymerase II-associated factor 1 homolog;PAF1 |
| Q7L266 | 8(12.7%) | 7(33.3%) | Isoaspartyl peptidase/L-asparaginase;ASRGL1 |

**Table 2.** 7 most frequent genes unique to cSCC samples, compared to BCC. Full table see **Table S4**

| GeneID | Frequency in samples # (%) | Frequency in patients # (%) | Gene Name |
| --- | --- | --- | --- |
| Q9GZP8 | 14(22.2%) | 10(47.6%) | Immortalization up-regulated protein;IMUP |
| F8WE70 | 17(27.0%) | 7(33.3%) | Serpin B13;SERPINB13 |
| P35658 | 13(20.6%) | 7(33.3%) | Nuclear pore complex protein Nup214;NUP214 |
| Q9UHR4 | 12(19.0%) | 7(33.3%) | Brain-specific angiogenesis inhibitor 1-associated protein 2-like protein 1;BAIAP2L1 |
| MAP4K4 | 11(17.5%) | 7(33.3%) | Mitogen-activated protein kinase kinase kinase 4;MAP4K4 |
| P13688 | 10(15.9%) | 7(33.3%) | Carcinoembryonic antigen-related cell adhesion molecule 1;CEACAM1 |
| ZNF827 | 9(14.3%) | 7(33.3%) | Zinc finger protein 827; Zinc finger protein 827 (Fragment);ZNF827 |

To identify most differentially expressed proteins we performed both parametric (Student T-Test, **Table S5**) and non-parametric (Wilcoxon rank-sum, **Table S6**) tests between cSCC and BCC samples. Both approaches identified a high number of significantly differentiated genes (978 and 944 overexpressed in BCC for p-value< 0.05 cut for T-Test and Wilcoxon rank-sum tests respectively). Interestingly, the opposite direction – overexpression in cSCC resulted in significantly less significant genes (378 and 125 overexpressed in cSCC for p-value< 0.05 cut for T-Test and Wilcoxon rank-sum tests respectively).

### Gene Ontology analysis of proteome extracted with e-biopsy from skin tumors

All 7087 identified proteins were sorted according to the lowest of two p-values (Student T-Test and Wilcoxon rank-sum, **Table S7, Table S8**) and fed into Gene Ontology (GO) analysis in terms of cellular processes, functions, and components with GOrilla tool^38–40^. We identified 31 cellular processes (**Table S9, Figure S2**), 23 cellular functions (**Table S10, Figure S3**) and 10 cellular components (**Table S11, Figure S4**) significantly (p-value<1e-03) enriched in the top of the list.

### Identification of protein signatures that differentiate cSCC and BCC lesions based on e-biopsy-sampled proteome

To identify a group of proteins with a maximal potential in distinguishing between cSCC and BCC conditions, we performed 100 repetitions of leave-6-patients out procedure (**Figure 4**). In each repetition, 18 cSCC and 18 BCC patients were randomly selected, and their proteomes were used to construct an optimal subset of 5 to 100 most differentiating proteins. The samples from the remaining 3 cSCC and 3 BCC patients were used to assess the classification quality of the resulted protein subset. Finally, for each protein we counted the number of repetitions it was selected for the final set of top-5 most differentiating proteins. Most frequently selected proteins were further studied for known connections and existing functions.

**Figure 4.**
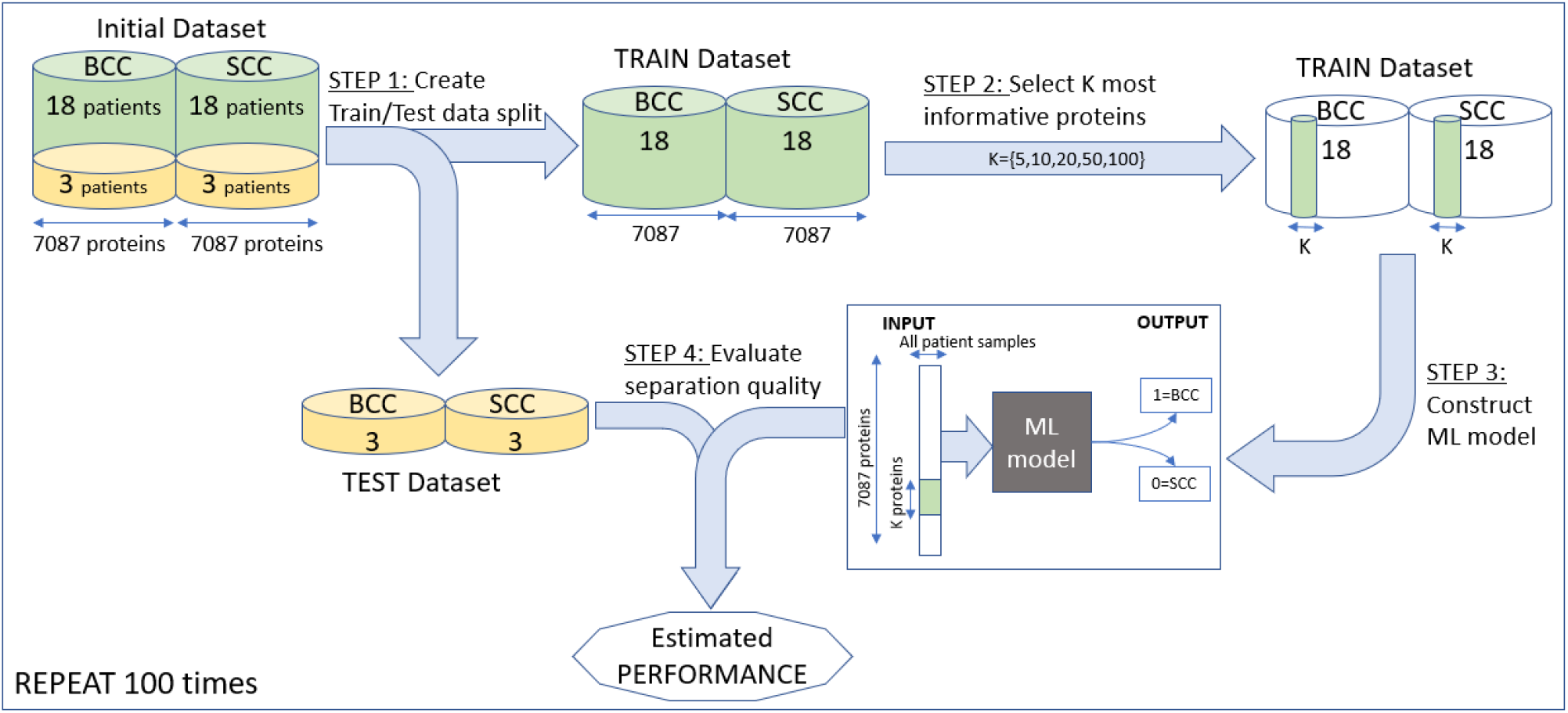
General flow of the leave-6-patient out algorithm for the selection of most informative proteins: (i) Splitting the data; (ii) Selecting most informative subset of proteins; (iii) Construction of Machine Learning model based on the selected subset; (iv) Evaluation of the classification performance of the resulted ML model.

Resulting average (over 100 repetitions) model accuracy ranges 74.0-81.0% (**Table 3, Table S12**). Addressing the cSCC subjects as positive and BCC subjects as negative, we achieved the positive predictive value (PPV) and sensitivity of 72.9-78.7% and 84.7-95.7% respectively, together with negative predictive value (NPV) and specificity of 80.1-90.3% and 60.0-69.7% respectively (**Table S12**). These findings are comparable to the quality of the initial manual diagnostics of the same patients in the clinics (**Table 4**), as performed by physicians, suggesting that the proposed molecular profiling has a potential to be a valuable diagnosis-supporting tool, especially if the large number of patients is used for model construction

**Table 3.** Performance of the best Machine Learning models (selected according to best leave-6-patient out accuracy). Per-model details are available in **Table S12**. All metrics are calculated over 100 repetitions and presented both as MEAN ± SEM (Standard Error of Mean) and as Clopper-Pearson 95% confidence interval (in parenthesis)

| Number of used proteins | Accuracy<br>(correct SCC and BCC predictions out of total cases) | NPV<br>(actual BCC out of predicted BCC) | Specificity<br>(predicted BCC out of total actual BCC) | PPV<br>(actual SCC out of predicted SCC) | Sensitivity<br>(predicted SCC out of total actual SCC) |
| --- | --- | --- | --- | --- | --- |
| 5 | 78.3% ± 1.6%<br>(74.8%-81.6%) | 84.2% ± 2.8%<br>(81.9%-91.0%) | 66.7% ± 2.9%<br>(61.0%-72.0%) | 76.2% ± 1.9%<br>(68.1%-77.4%) | 90.0% ± 1.8%<br>(86.0%-93.2%) |
| 10 | 81.0% ± 1.7%<br>(77.6%-84.1%) | 86.6% ± 2.7%<br>(85.5%-93.6%) | 69.7% ± 2.9%<br>(64.1%-74.8%) | 78.7% ± 1.8%<br>(70.5%-79.6%) | 92.3% ± 1.5%<br>(88.7%-95.1%) |
| 20 | 79.5% ± 1.6%<br>(76.0%-82.7%) | 88.0% ± 2.5%<br>(84.5%-93.0%) | 67.0% ± 2.7%<br>(61.4%-72.3%) | 76.5% ± 1.7%<br>(68.8%-78.0%) | 92.0% ± 1.6%<br>(88.3%-94.8%) |
| 50 | 79.2% ± 1.7%<br>(75.7%-82.3%) | 90.3% ± 2.6%<br>(89.2%-96.5%) | 62.7% ± 2.9%<br>(56.9%-68.2%) | 75.0% ± 1.7%<br>(67.2%-76.3%) | 95.7% ± 1.2%<br>(92.7%-97.7%) |
| 100 | 78.5% ± 1.7%<br>(75.0%-81.7%) | 88.9% ± 2.8%<br>(88.0%-95.8%) | 62.0% ± 2.8%<br>(56.2%-67.5%) | 74.1% ± 1.7%<br>(66.7%-75.8%) | 95.0% ± 1.4%<br>(91.9%-97.2%) |

**Table 4.**
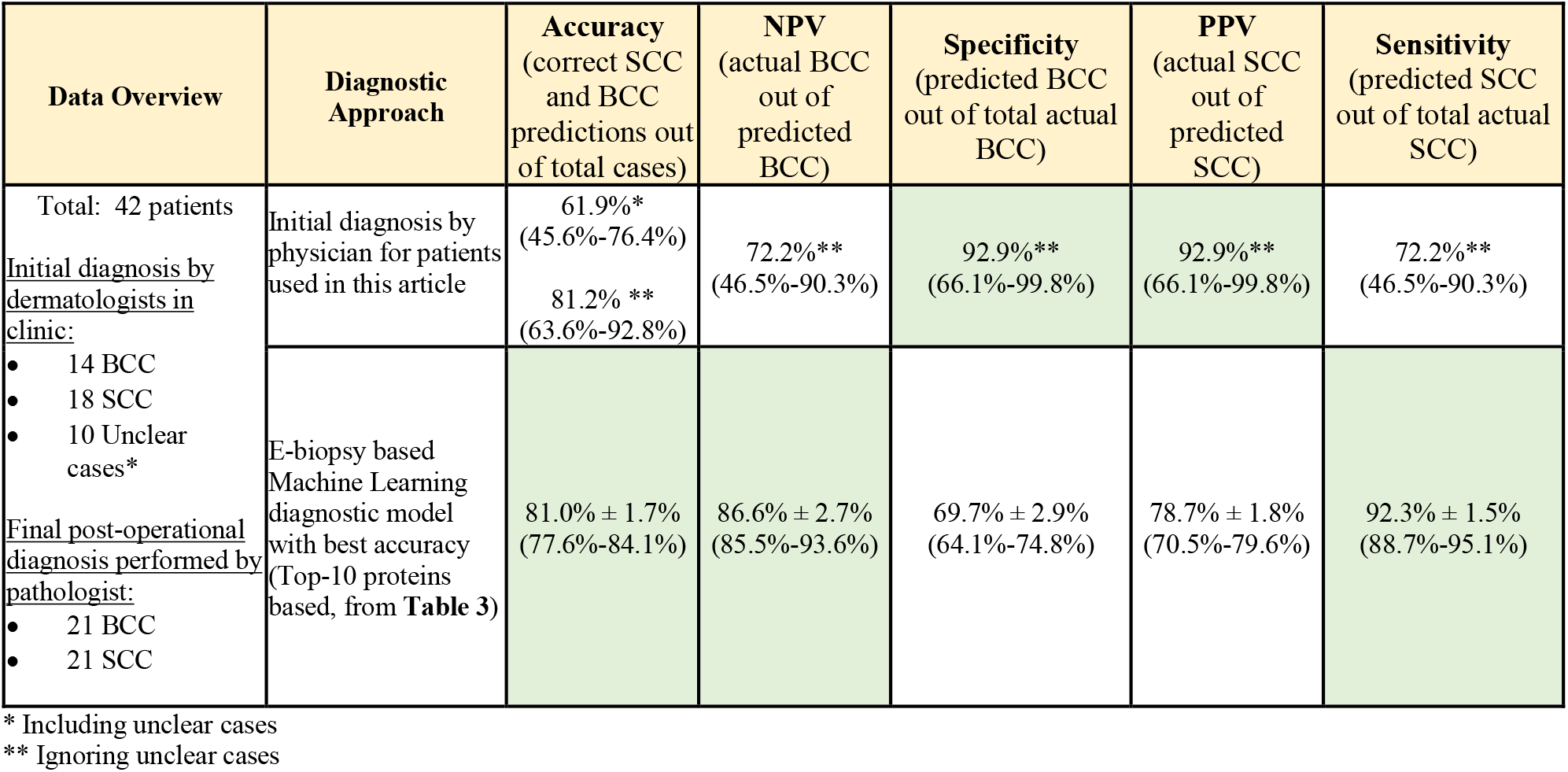
Comparison of the best-performing Machine Learning model with accuracy of the initial diagnosis in clinics, performed by physicians. In parenthesis we present Clopper-Pearson 95% confidence interval for each metric. Cells with the best observed performance are highlighted.

### Differentially expressed proteins and a protein network that differentiates cSCC from BCC

Summarizing the genes selected as top-5 most useful for constructing the machine learning classifiers between BCC and cSCC over different repetitions of the leave-6-patient out process (**Figure 4**), we derived the following 11 genes presented in **Table 5, Figure 5**. Most of these proteins are known to be associated with malignancies, including BCC or cSCC ^42^.

**Table 5.** Genes most frequently selected in the final subset of top-5 genes differentiating cSCC and BCC.

| Gene Symbol | Gene Uniprot Id | Gene Name | Direction in cSCC compared to BCC | Number of repetitions the gene was selected into final Top-5 |
| --- | --- | --- | --- | --- |
| CRNN | Q9UBG3 | Cornulin | DOWN | 92 / 100 |
| SULT1E1 | P49888 | Sulfotransferase 1E1 | DOWN | 65 / 100 |
| H1-5 | P16401 | Histone H1.5 | DOWN | 41 / 100 |
| ISG15 | P05161 | Ubiquitin-like protein ISG15 | UP | 27 / 100 |
| ERO1A | Q96HE7 | ERO1-like protein alpha | UP | 26 / 100 |
| KRT27 | Q7Z3Y8 | Keratin, type I cytoskeletal 27 | DOWN | 26 / 100 |
| KATNAL2 | Q8IYT4 | Katanin p60 ATPase-containing subunit A-like 2 | DOWN | 22 / 100 |
| PALM | O75781 | Paralemmin-1 | DOWN | 22 / 100 |
| VCAN* | P13611* | Versican core protein* | DOWN | 19 / 100 |
| HNRNPDL | O14979 | Heterogeneous nuclear ribonucleoprotein D-like | DOWN | 13 / 100 |
| KYNU | Q16719 | Kynureninase | UP | 11 / 100 |

\* VCAN does not appear in **Figure 6**

**Figure 5.**
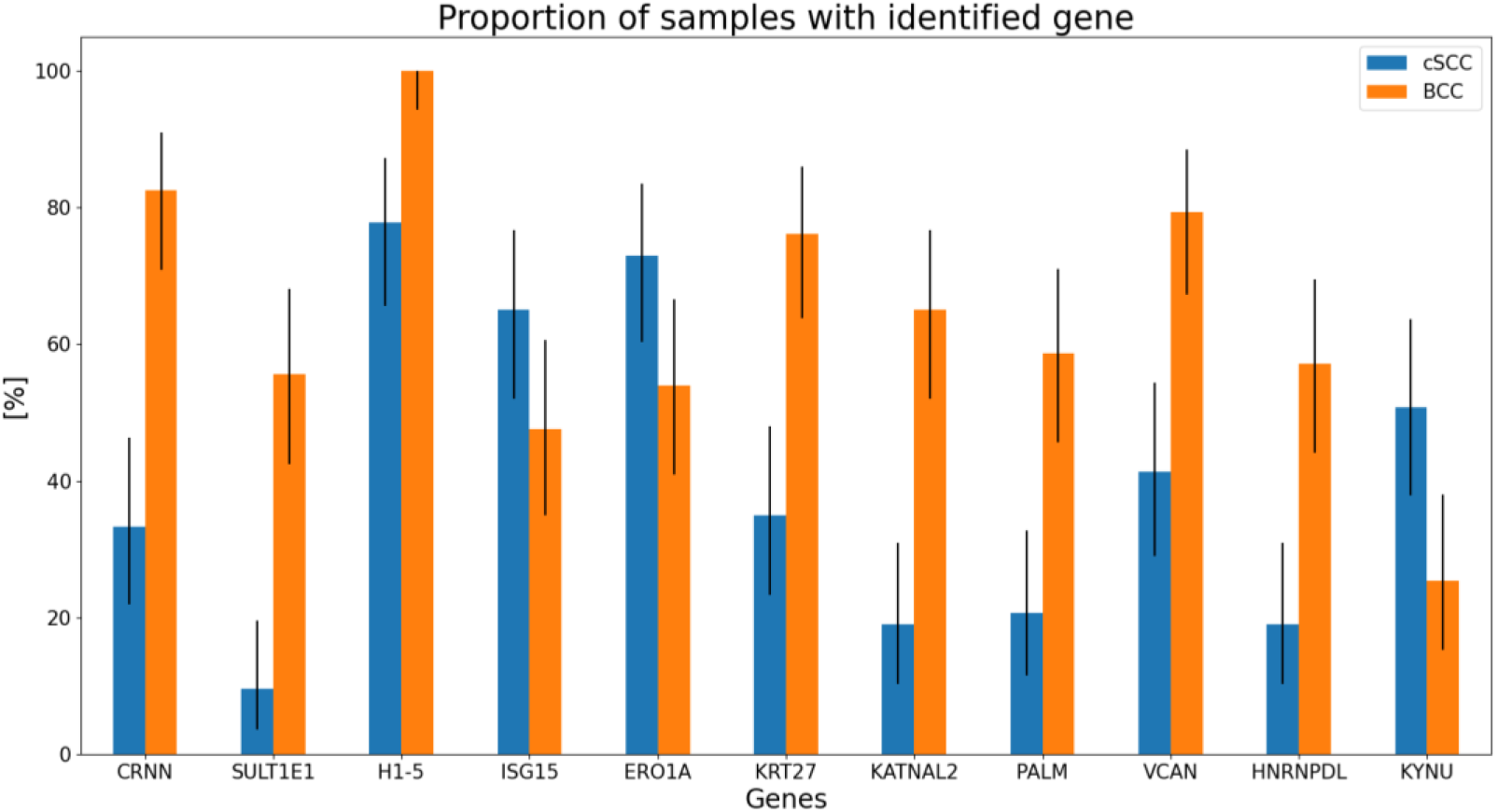
Comparative proportion of samples, where the genes most optimally differentiating between two cancers from **Table 5** were identified. Whiskers depict 95% Clopper-Pearson confidence interval.

Moreover, protein-protein interaction data from IntAct DB^43^ together with pathways derived from KEGG DB^44^ and other public sources^45,46^ (**Table S13, Table S14**) provides an interesting protein-protein interaction map for most of these proteins, with the exception of VCAN (**Figure 6**).

**Figure 6.**
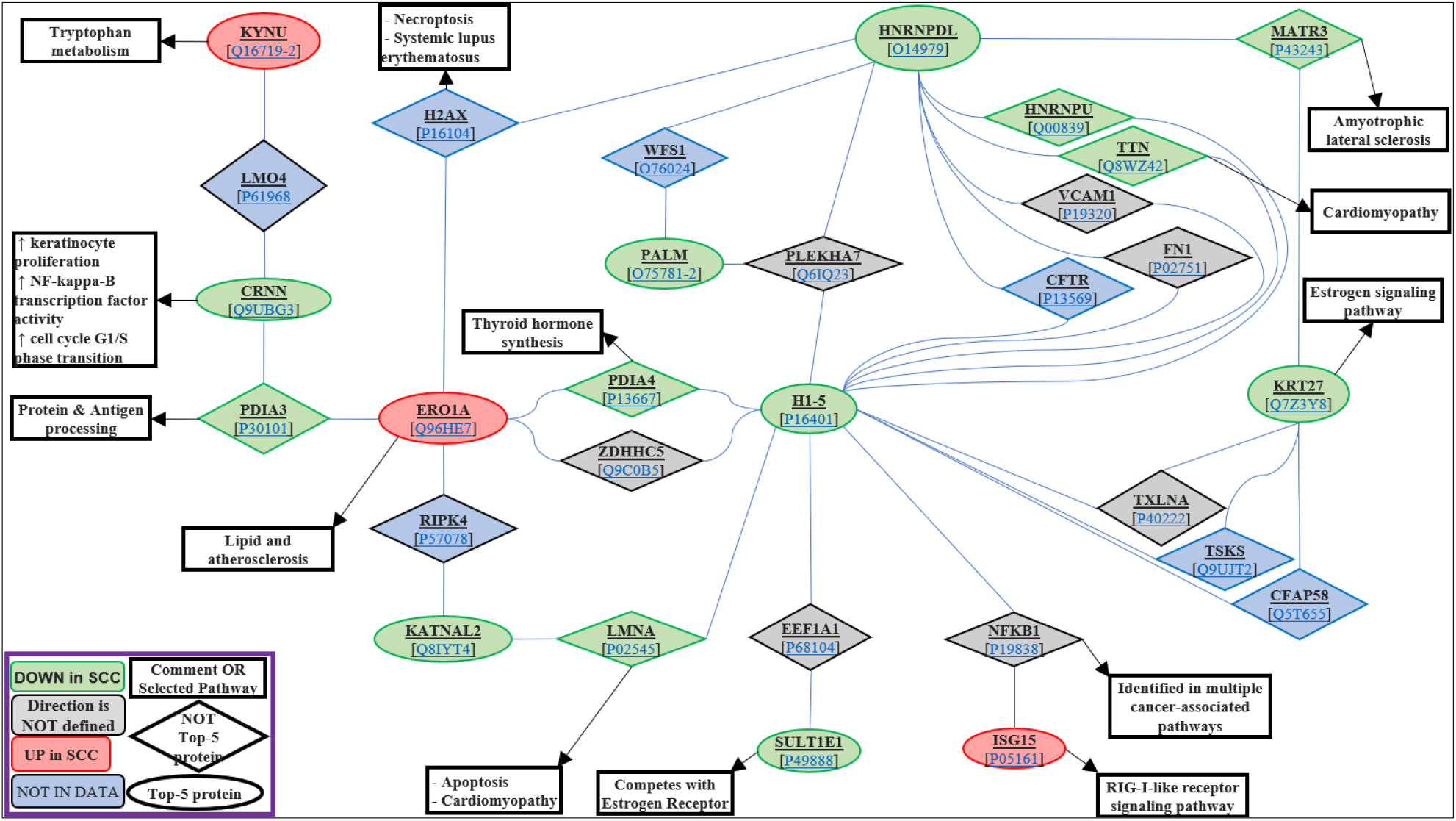
Protein interaction network of genes most frequently selected to the top-5 optimal differentiating gene subset (**Table 5**) based on IntAct DB^43^. More details on the gene appearance, PPI details and known pathways are in **Table S13** and **Table S14**. Shape notation: ellipses correspond to proteins from **Table 5**; rhombs correspond to proteins connecting between them; rectangles correspond to pathways and comments. Shape color: green corresponds to proteins with intensity in cSCC smaller than in BCC; red corresponds to proteins with intensity in cSCC greater than in BCC; gray corresponds to proteins without significant change in measured intensity in cSCC vs BCC; blue corresponds to proteins that were not observed in the data.

## Discussion

Current solutions for diagnosing skin tumors lack the ability to sample suspicious lesions for molecular content without resection or thermal destruction. In this work we introduced the potential of e-biopsy, a novel method of molecular sampling with electroporation, for applications in molecular diagnostics of non-melanoma skin cancers. Specifically, we showed that proteome released by electroporation and directly harvested into a syringe by vacuum, has a signature that differentiates cSCC from BCC. We analyzed the sampled proteomes to identify proteins uniquely expressed in each type of the tumors, and performed differential expression analysis followed by GO term enrichment analysis and protein-protein interaction analysis. We also developed a classifier to differentiate cSCC from BCC with a performance comparable to that of human medical professionals. Finally, we modelled electric and thermal effects of the e-biopsy procedure on the excised human skin and evaluated the e-biopsy technique reproducibility in terms of the harvested proteomic samples.

In this study we found that e-biopsy sampled proteome showed 17 proteins uniquely expressed in BCC (**Table 1**) and 7 proteins uniquely expressed in cSCC (**Table 2**). Furthermore, 11 genes (**Table 5, Figure 5**) were identified as most informative for the optimal separation between the two cancers. Unfortunately, none of these 11 genes appeared unique to the specific carcinoma type (**Figure 5**), thus the optimal differentiation of cSCC from BCC would require joint measurements of several genes.

Cornulin, CRNN gene (selected by 92% of the machine learning models, **Table 5, Figure 5**), downregulated in cSCC in comparison to BCC, plays a role in epidermal differentiation, while its expression is believed to be specific to squamous cells^47^. It was shown previously that CRNN is downregulated in tongue SCC^48,49^. In addition, CRNN expression has been reported to differentiate between low-grade and high-grade oral epithelial dysplasia and could be represented as a potential biomarker for the assessment of progression of oral cancers^50,51^. Up-regulated CRNN levels prevent lesion formation, and its tumor suppressive role has been reported^51^, which is aligned with the less aggressive nature of BCC. Sulfotransferase 1E1 (SULT1E1), which also appears in BCC with increased measured intensity in comparison to cSCC (**Table 5, Figure 5**), is responsible for the metabolism of active estrogens and plays crucial roles in their homeostasis^52^. SULT1E1 was reported as a predictor of breast cancer recurrence^53^ and its upregulation has been reported in normal mammary epithelial cells and breast cancer cell lines^54–56^. SULT1E1 downregulation was proposed to serve as a marker for more aggressive cancer^57^. Ubiquitin-like protein Interferon Stimulated Gene (ISG15)^58^, upregulated in cSCC in our study in comparison to BCC, was overexpressed in up to 80% of oral squamous cell carcinoma as reported in previous studies^59^. The endoplasmic reticulum oxidoreductin-1-like (ERO1-like protein) alpha, upregulated in cSCC in comparison to BCC as sampled by e-biopsy proteomes (**Table 5, Figure 5**) was shown in literature to be associated with cancer^60^, driving the production of VEGF^61^. Interestingly, another study showed significantly higher peritumoral and intra-tumoral blood vessel area in cSCC when compared to BCC^62^. Also, versican core protein was previously shown to be a mediator of skin cancer development in mice and humans and was reported to be strongly expressed in BCC compared to cSCC^42^ similar to our findings (**Table 5, Figure 5**). Finally, the protein-protein interaction analysis of these 11 highly informative genes (**Table 5, Figure 5**) enabled us to identify a novel protein interaction network (**Figure 6**), which surprisingly covers 10 of the 11 top-ranked proteins. Such network has the potential to be valuable for further understanding of the differences in molecular mechanisms of the two carcinomas. We report, in this study, a binary classifier differentiating between cSCC and BCC based on a limited, predetermined number of proteins. A subset of 5 to 100 proteins resulted in a classifier accuracy of 78.3-81.0% (depending on the number of proteins used, **Table 3**). In comparison, the accuracy of the initial diagnosis by dermatologists in clinic on the same 42 patients was only 61.9% (or 81.2% when omitting the unclear cases, **Table 4**). However, these 42 patients selected for our study do not represent the general distribution of patients in clinics. For example, an expert panel of pathologists reported on 91.6% accuracy for 154 patient panel ^13^, and a single surgeon reported on accuracy of 93.8% in 1326 patient cases ^41^. It is important to notice that both the panel and surgeon reports included significantly higher number of BCC patients in comparison with cSCC, while in our study the numbers of cSCC and BCC patients are equal.

One of the limitations of the current study is its sole focus on the NMSC samples, without addressing its matching healthy areas. Thus, for the next stage we are working on gathering healthy tissue samples and their comparative analysis to the available NMSC samples. Moreover, we plan to include paired healthy and affected samples obtained from the same patient to verify that every patient’s lesion has a genetically adequate control. Based on proteomic signatures obtained by e-biopsy and the resulted model performance, we aim to develop even better classifiers differentiating cSCC, BCC and normal skin in future large-scale trials. Such disease-specific classifiers could enable a personalized treatment approach in high risk NMSC.

Additional limitation of this study is that all sampled proteomes were measured using LC-MS/MS. Thus, measurement is biased towards this method. Therefore, we are working on a targeted proteomics confirmation study to validate the presence or absence of the selected proteins. One of the most problematic side effects of high voltage pulsed electric fields application on skin is Joule heating^63^. High temperature may have devastating effects on living cells, such as protein coagulation, microvascular blood flow stasis, cell death or permanent changes in tissue structure or function. When the temperature is over 42°C for a prolonged exposure time, thermal damage begins. The results of our models show that under current parameters (*ex-vivo*), the procedure is non-thermal. Future *in vivo* applications will require more detailed thermal modeling and testing^64–66^, once the geometry of the applicators is known. The decision to biopsy the lesion on the skin should be made only if the biopsy would be helpful to the diagnosis and would provide the information needed to the clinician. The major types of biopsies currently used include punch biopsy, shave biopsy, excision biopsy, and curettage biopsies. Our proposed e-biopsy technique could result in a novel sampling device and approach enabling personalized medicine by providing a minimally invasive local molecular sampling procedure to assist in deciding whether tissue biopsy is needed. In addition, e-biopsy could assist in the personalization of skin lesion care in the future, potentially reducing morbidity and mortality by enhancing and speeding-up the decision process on the use of more advanced surgeries or adjuvant therapies such as: radiation, chemotherapy, and emerging immunotherapy. E-biopsy derived results can also contribute to determining the recommended frequency of follow up visits and dictate the type and frequency of imaging needed for follow up. Until this moment, there are no known approved biomarkers to distinguish high risk NMSC from the benign subtype. We envision that *in vivo* sampling of molecular signatures with e-biopsy, such as proposed for development in this work, will enable future classification of NMSC molecular signatures to groups by observed multi-omics profiles and tumor phenotypes, such as aggressiveness (defined by local recurrence, lymph node metastasis, systemic metastasis). These group characteristics together with patient’s demographics (gender, age, etc.) and clinical information (immunosuppression status, used medications, etc.) pave the way to enable personalized diagnostics and more precise treatments, improved outcomes and savings in costs and resources.

## Materials and Methods

From March 2020 to March 2022, tissue samples were collected from NMSC (non-melanoma skin cancer) lesions from 143 patients who underwent surgical excision of a skin lesion suspected as BCC and cSCC, at Meir Medical Center, Israel. All lesions excised were at least 1 cm in diameter. Proteins were extracted from 42 patients fresh (between 10-20minutes after surgery) samples randomly chosen 21 c BCC and 21 from the entire patent’s data (126 samples, 3 per tumor, one tumor per patient) and proteomics were analyzed as described below. This study was approved by the Meir Medical Center IRB, number MMC-19-0230. All patients gave consent for participation and for genetic analysis of tissue.

### Sample extraction with e-biopsy

Each patient’s lesion was sampled in 3 locations, resulting in 63 samples for each carcinoma type. The sampling needle is inserted in the sampling location and the ground needle is positioned on the skin surface without the penetration and the PEF are applied. A vacuum is applied on the same needle through which the PEF pulses are delivered, to pump the released cellular content into the needle and the syringe. The pulsed electric field was applied using our laboratory custom-made high-voltage pulsed electric field generator, described in detail in ref^67^. E-biopsy was performed using a combination of high-voltage short pulses with low-voltage long pulses^24,68^ as follows: 40 pulses, 1000 V, 40 µs, 4 Hz, and 40 pulses, 50 V, 5 ms, delivered at 4 Hz. After the application of the electric fields, the liquids were extracted from the tissue to the needle with the vacuum, manually applied with a 1.5mL syringe. The liquids were immediately transferred to 1.5ml tubes with 100µl double distilled water. The electrodes were positions about 5mm apart. For the sampling electrode we used a standard 30-G insulin syringe. The second electrode was a custom-made bar with 3mm diameter.

### Proteins isolation from the e-biopsy sample

Proteins were isolated from the e-biopsy extract using the EZ-RNA II kit (Biological Industries, Beit Haemek Ltd). Homogenizing solutions were not used in the samples; phase separation solutions were directly added as follows: 0.2 ml of water-saturated phenol, and 0.045 ml of BCP. This step was followed by protein precipitation using isopropanol and wash using guanidine hydrochloride in 95% ethanol. Airdried protein pellets were taken for proteomic analysis as described below.

### Identifying and quantifying proteins with LC-MS/MS Proteolysis

The samples were brought to 8M urea, 400mM ammonium bicarbonate, 10mM DTT, vortexed, sonicated for 5’ at 90% with 10-10 cycles, and centrifuged. The protein amount was estimated using Bradford readings. 20ug protein from each sample was reduced 60ºC for 30 min, modified with 37.5mM iodoacetamide in 400mM ammonium bicarbonate (in the dark, room temperature for 30 min) and digested in 2M Urea, 100mM ammonium bicarbonate with modified trypsin (Promega) at a 1:50 enzyme-to-substrate ratio, overnight at 37°C. Additional second digestion with trypsin was done for 4 hours at 37°C.

### Mass spectrometry analysis

The tryptic peptides were desalted using C18 tips (Harvard Apparatus,MA), dried, and re-suspended in 0.1% formic acid. The peptides were resolved by reverse-phase chromatography on 0.075 × 180-mm fused silica capillaries (J&W) packed with Reprosil reversed-phase material (Dr. Maisch GmbH, Germany). The peptides were eluted with a linear 180-minute gradient of 5 to 28%, 15 minutes gradient of 28 to 95%, and 25 minutes at 95% acetonitrile with 0.1% formic acid in water at flow rates of 0.15 μl/min. Mass spectrometry was performed using Q-Exactive Plus mass spectrometer (Thermo Fischer Scientific, CA) in a positive mode using a repetitively full MS scan followed by collision-induced dissociation (HCD) of the 10 most dominant ions selected from the first MS scan. The mass spectrometry data from all the biological repeats were analyzed using the MaxQuant software 1.5.2.8 vs. the human proteome from the UniProt database with 1% FDR. The data were quantified by label-free analysis using the same software, based on extracted ion currents (XICs) of peptides enabling quantitation from each LC/MS/MS run for each peptide identified in any of the experiments.

### Bioinformatics data preprocessing and identification of unique proteins

Raw mass spectrometry data received from the MaxQuant program was transformed into a binary format. Specifically, a protein value measured in the sample was assigned with 1’ if it was observed in this sample with any positive intensity and assigned with 0’ otherwise. Proteins were defined unique for the certain condition, if they were observed only among samples with this condition.

### Differential Expression and GO terms analyses

Protein differential expression analysis between cSCC and BCC was performed both with a parametric Student TTest (*scipy*.*stats*.*ttest_ind*) and with a non-parametric Wilcoxon rank-sum (*scipy*.*stats*.*ranksums*) tests. Afterwards, for each protein the lowest (among received two) test p-value was selected and corrected using Bonferroni approach with multiplication by 2. Next, the proteins were sorted according to the resulted p-values and fed into Gene Ontology (GO) analysis with GOrilla tool^38–40^ (exact input is available in **Table S8**) to identify statistically significant overrepresentations of specific cellular processes, functions, and components.

### Constructing protein signatures differentiating cSCC and BCC

Protein signature differentiating cSCC from BCC is effectively a small set of proteins that, when measured together, is informative enough to separate between these two conditions. Selection of these most informative proteins was performed by 100 repetitions of leave-6-patient out algorithm, while in each repetition 18 BCC and 18 SCC patients were used as a training set for protein selection and the remaining 3 BCC and 3 SCC patients were used as a testing set to assess the performance of the resulting set of proteins.

In each repetition of the leave-6-patient out the training part consisted of two major stages: (i) verification of high gene abundance among cSCC or BCC training samples; followed by (ii) iterative backward gene elimination based on the relative protein importance. In the first stage, we omitted all the proteins with maximal prevalence *below 80%* among 18 BCC and 18 cSCC training patients and with maximal prevalence *below 50%* among 54 BCC and 54 cSCC training samples (each patient was sampled in 3 locations). With this procedure we aim to increase the chances to observe the selected marker in the sample of the corresponding carcinoma type. On average (over different repetitions), roughly 1700 out of 7087 proteins passed this stage in each repetition. In the second stage, we iteratively omitted 10% of the least important proteins, where the *importance* of each protein was defined as average protein importance obtained from 3 different machine learning classifiers: (i) Random Forest (RF); (ii) Logistic Regression (LR) and (iii) Deep Neural Network (DNN). Specifically we used: (i) rank of feature importance for the corresponding protein as obtained from *sklearn* Random Forest classifier; (ii) rank of absolute coefficient for the corresponding protein as obtained from Logistic Regression classifier; and (iii) rank of total absolute weight of the input feature for the corresponding gene in the input layer of Deep Neural Network classifier. The predicted per-patient diagnosis (cSCC patients were addressed as positive and BCC patients as negative) was estimated as average over predictions of 3 patient’s samples. The resulted model performance was estimated in terms of overall accuracy, positive predictive value (PPV), negative predictive value (NPV), sensitivity and specificity over the 6 patients from the testing set.

Finally, all the metrics were summarized in terms of mean and its standard error (standard error of mean (SEM) is defined as standard deviation (STD) divided by the square root of the sample size) over 100 repetitions and 95% Clopper-Pearson confidence interval for the resulted value was estimated.

### Analysis of most differentiating proteins

The final set of most differentiating proteins was derived from the proteins that were consistently (over 100 repetitions of leave-6-patients-out feature selection algorithm) selected into the final set of top-5 genes. Specifically, we identified 11 proteins that were chosen with at least 10% frequency (10 reps or more; **Table 5**).

These proteins were analyzed for protein-protein interactions based on IntAct DB^43^ resulting in a protein network describing these and intermediate proteins. This network, together with protein differential expression directions and with known selected pathways derived from KEGG DB^44^ and other public sources^45,46^, is presented in **Figure 6**, while it’s exact details including (i) protein abundances in our patients, (ii) details and references for experiments that observed each protein-protein interaction, and (iii) known pathways for each protein are presented in **Table S10** and **Table S11**.

### Numerical simulations of electric fields distribution in the skin tissue and electric field-induced thermal effects

To model the distribution of the electric fields in the skin with tumors during e-biopsy, we used the finite elements method (FEM), which allows us to find an approximate solution in complex geometries for solving the Laplace differentiation equation with boundary conditions defined by the applied voltage. Numerical solutions for a Laplace equation that result in the electric field distribution in the brain and brain melanoma models were performed in QuickField (Terra Analysis, Denmark). The electric and thermal properties of tissues used to appear in **Table S1**. The model files with full solutions appear online at the following link: https://github.com/GolbergLab/BCCvsSCC.git

We assume the thermal properties of the skin didn’t change after electroporation^69^, while the electric conductivity after electroporation increased ^70^. Direct current (DC) conduction and steady-state heat transfer problems were coupled with transient heat field problems.

In steady-state heat transfer, with Dirichlet boundary conditions, the temperature is constant with time: TAirline = 25 °C, where the airline differentiates between skin and air.

Heat sources were imported from DC conduction and steady-state heat transfer problems coupling, for the thermal field problem. To calculate the power supplied by the pulsed electric field, we used the following equation (**Eq. 1**):

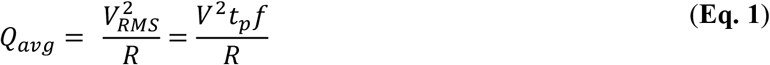

where *Q*_*avg*_*(W)* is the total average power delivered by square pulse electric field, R (ohm) is the resistance, *V*_*RMS*_ is the root mean square voltage, *V (Volt)* is the applied voltage, *t*_*p*_ is the duration of the pulse and *f (Hz)* is the frequency of the pulse wave.

To calculate the electric field distribution, we used the Laplace equation (**Eq. 2**):

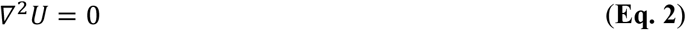

With the following potentials: *V*_*Short, high voltage pulse*_ = 1000*V, V*_*Long, low voltage pulse*_ = 50*V*, and *V*_*Ground*_ = 0.

To calculate the thermal distribution, we solved the transient heat transfer equation (**Eq. 3**):

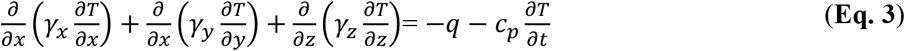

Where *T* is the temperature (*K*), *γ*(W *K*^−1^*m*^−1^) is the thermal conductivity, *c*_*p*_ (*J K*^−1^ *kg*^−1^) is the specific heat capacitance, *t* (s) is time, *q* (*Wm*^−3^) is the volume power of heat sources. In our problem *q* is the average volume power supplied by a pulsed electric field. We assume that heat is transferred by convection between the air, and skin, and the convection coefficient with air is *α =* 5*(W K*^*-1*^*m*^*-2*^)^71^

### Reproducibility analysis

To assess the reproducibility of e-biopsy methodology, the similarity between the measurements gathered from 3 sampled patient’s locations was estimated. Our assumption is that the actual proteomes in the sampled locations should be very similar, given these locations are spatially and phenotypically close. Therefore, we expect that protein profiles sampled by a reliable technology to be in a high agreement with each other. Together with this, inherent tissue spatial heterogeneity would prevent even from the ideal sampling method to receive the exact measurement replicas.

Specifically, to assess the similarity of the samples produced by e-biopsy method, while reducing the impact of local spatial heterogeneity, we calculated maximal intra-patient Pearson (*scipy*.*stats*.*pearsonr*) correlation between each of 3 pairs of measured raw protein intensities. The per-patient results are available in **Table S2**.

### Calculating FDR for condition-unique proteins

To assess the FDR for the number of proteins identified as *unique* in a certain condition, we calculated the probability to observe a protein uniquely in this condition for the predefined number of times (or more) by a mere chance. Specifically, the probability of a certain protein *t* to appear in at least 4 (and at most 63) BCC samples, while to never appear in any of 63 cSCC samples (and symmetrically vice versa, since the data is balanced) by a mere chance was 3.2e-03. This is derived as follows (**Eq. 4**), where *HG(126,k,63,k)* is a hypergeometric probability of selecting *k* out of *k* samples inside the subgroup sized 63 in population sized 126; and *P*(***t*** occurs in exactly ***k*** cases) is calculated from data:

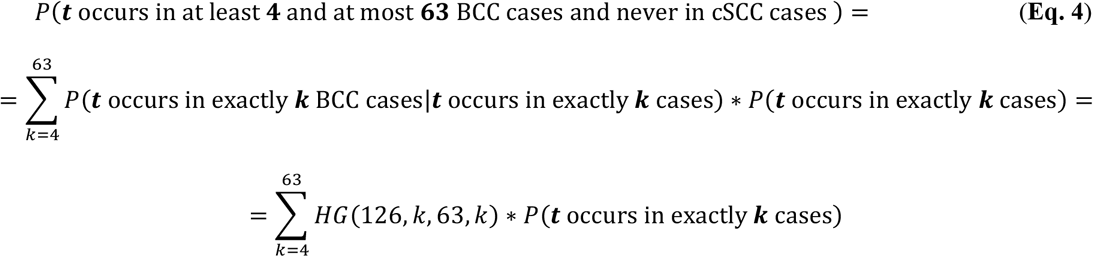

This leads to the expectation of 22.9 such proteins, resulting in FDR of 1.94e-01 for 118 such proteins uniquely observed in BCC and of 2.29e-01 for 100 such proteins uniquely observed in cSCC samples. On the patient level, the probability of a certain protein to appear in at least 7 (and at most 21) BCC patients, while to never appear in any of 21 cSCC patients (and symmetrically vice versa) by a mere chance is 2.3e-04 (calculation is similar to **Eq. 4**), leading to the expectation of 1.7 such proteins. This results in FDR of 9.79e-02 for 17 such proteins uniquely observed in BCC and of 2.38e-01 for 7 such proteins uniquely observed in cSCC patients.

## Supporting information

Supplementary Content and link to GitHub repository

## Data Availability

All data produced in the present study are available upon reasonable request to the authors

https://github.com/GolbergLab/BCCvsSCC

## Data availability

The authors hereby declare that all data supporting the findings of this study are available within the paper and its **Supplementary Information**.

## Authors contributions

Edward Vitkin – conceptualization, bioinformatics, manuscript drafting and approval

Julia Wise – experiments, protein sampling and analysis

Ariel Berl – experiments, samples collection, pathology, clinics, manuscript review

Ofir Shir-az – experiments, samples collection, pathology, clinics

Batel Gabay – numerical modeling, manuscript drafting

Amrita Singh – experiments, protein sampling and analysis

Vladimir Kravtsov – pathology

Zohar Yakhini – conceptualization, data analysis, critical manuscript review.

Avshalom Shalom – conceptualization, critical manuscript review

Alexander Golberg – conceptualization, experiment, data analysis, manuscript drafting.

## Acknowledgments

The authors thank the Israel Ministry of Science and Technology, Israel Innovation authority Kamin project, the TAU SPARK fund, TAU Zimin Center for technologies for better life and the EuroNanoMed MATISSE project for their support of this project. All authors thank the members of the Smoler Proteomics Center at the Faculty of Biology at the Technion. We specifically thank Keren Bendalak for her help with the LC-MS/MS analysis.

