## Supplementary Content and link to GitHub repository for "Proteome sampling with e-biopsy enables differentiation between cutaneous squamous cell carcinoma and basal cell carcinoma"

### Supplementary Information

https://github.com/GolbergLab/BCCvsSCC

**Table S1.** Electrical and thermal properties of human skin used for the numerical simulations.

**Table S2.** Reproducibility analysis of molecular harvesting with e-biopsy

**Table S3.** Sampled proteins unique to BCC

**Table S4.** Sampled proteins unique to cSCC

**Table S5.** Student TTest of SCC vs BCC

**Table S6.** Wilcoxon rank sum test of SCC vs BCC

**Table S7.** Differential expression summary of SCC vs BCC

**Table S8.** Input to GOrilla tool

**Table S9.** GOrilla Output. GO Processes significantly different between cSCC and BCC patients

**Table S10.** GOrilla Output. GO Functions significantly different between cSCC and BCC patients

**Table S11.** GOrilla Output. GO Components significantly different between cSCC and BCC patients

**Table S12.** Performances of different Machine Learning models over different amounts of selected proteins

**Table S13.** References (from IntAct DB) used to construct the protein-protein interaction map on **Figure 6.**

**Table S14.** Pathways where proteins from **Table 5** participate.

**Figure S1** Patient Demographics
**a.** Gender distribution of patients; **b.** Age distribution of patients; **c.** Country of birth distribution of patients

**Figure S2** GO Processes significantly different between cSCC and BCC patients

**Figure S3** GO Functions significantly different between cSCC and BCC patients

**Figure S4** GO Components significantly different between cSCC and BCC patients
